# Peripheral intravenous catheter fixation splint in pediatrics, FIX I/V: A proof-of-concept study protocol

**DOI:** 10.64898/2026.11.07.26367513

**Authors:** Sonia Dubé, Abir El Haouly

## Abstract

**Introduction:** Using a fixation splint to maintain a peripheral intravenous device (PIVD) is common practice in pediatrics. However, splints are made from small wooden, plastic, or metal boards, and wrapped with gauze and adhesive fabrics. Accidental removals of the PIVD, mechanical complications (infiltrations/extravasations, occlusions, etc.), and skin injuries related to the splint are frequently observed. They do not always keep the PIVD securely in place. In addition, painful procedures to reinstall the PIVD in the child can worsen the negative experience of hospitalization. Moreover, unplanned PIVDs and fixation splint reinstallation appears to increase nurse workload and, consequently, care costs. New PIVD fixation splints for the hand and arm, adapted for newborns, infants, and children, are available in Quebec: the FIX/IV splints. Field observations suggest these splints show promising safety and efficacy. A prospective interventional proof-of-concept study is being conducted.

**Objective:** This article describes the protocol for a prospective interventional proof-of-concept study on the safety and acceptability of the new pediatric FIX/IV PIVD fixation splints.

**Method:** This protocol describes the implementation process conducted in fall 2023 and winter 2024. We selected 30 children for the experimental group and 22 for the control group. We will conduct descriptive and comparative statistics between groups, as well as a questionnaire on the acceptability of these new splints among nurses and parents.

**Discussion and conclusion:** This proof-of-concept protocol presents the expected safety criteria for these new splints, which will improve the rigour and scientific value of future clinical trial phases with a larger sample.

The authors published this protocol after data collection but before analysis. Two interns collected the data, while the researchers will conduct the analysis and publish the results.

## Background et rationale

For many hospitalized children, placing a PIVD is necessary to administer medications, fluids and other biological products. This invasive and painful procedure is the most frequent in hospitals due to repeated failures and accidental or premature removal of the PIVD (Legemaat & al., 2016; Rozsa & al., 2015). Studies estimate the failure and malfunction rate of a PIVD to be between 36% and 55%, and the rate of mechanical complications, such as occlusions, infiltrations, extravasations, phlebitis and infections, between 25% and 67% (Chen YM. & al., 2022; Legemaat & al., 2016; Malyon & al., 2014; Marsh & al., 2024). To maintain PIVD placement, clinicians commonly use fixation splints in pediatrics and neonatology. These devices are frequently used in young children due to agitation and the difficulty of immobilizing joints near the catheter insertion site, particularly the wrist and elbow (Harris et al., 2022; Huang et al., 2021). Their purpose is to immobilize the joint near the insertion site to protect the venous access, extend its lifespan, and prevent mechanical complications as well as premature or accidental removal (Harris & al., 2022; Serane V & al., 2022).

On care units, conventional fixation splints for a PIVD are made from a wooden, plastic, or metal board wrapped in gauze. These splints are secured to the limb using gauze bandages and adhesive tape. When using a conventional splint, special attention must be given to keeping the PIVD insertion site visible to allow for continuous clinical monitoring. Furthermore, conventional splints do not always provide secure stabilization, as they cover only part of the arm or hand, and the gauze bandages and adhesive tape are often insufficient to maintain stability throughout treatment. Consequently, this can lead to mechanical complications, skin injuries, and the premature or accidental removal of the PIVD (Sweeney & al., 2018). Such circumstances require the nursing staff to reinsert and re-secure the PIVD, sometimes multiple times. This ultimately increases painful episodes, anxiety for both the child and their parents, and costs related to clinical intervention time (Helm et al., 2015; Schmutz & al., 2020).

### Issues related to the safety and effectiveness of splints

Studies evaluating the safety and effectiveness of fixation splints for PIVD in pediatrics and neonatology are scarce, if not rare. For example, Serane & al. (2022) documented the effect of conventional fixation splints on PIVD lifespan in neonatology. Based on a randomized sample of 159 premature newborns, they concluded that these splints did not prolong the intended lifespan of the PIVD for treatment and could cause skin injuries. Harris & al. (2022), for their part, evaluated the safety, effectiveness and acceptability of a novel, non-conventional PIVD fixation splint -which does not require adhesive tape or bandaging on the skin - on 38 premature newborns in neonatology. They highlighted that these splints were more effective at maintaining PIVD stability and reduced skin injuries compared to conventional splints. Due to the scarcity of data, these results remain preliminary and do not allow for generalizable conclusions (Sweeney & al., 2018). Moreover, to our knowledge, no study has evaluated the effectiveness and safety of PIVD fixation splints in pediatrics. Nonetheless, the high failure rate of PIVD and their fixation mechanisms raises safety and effectiveness concerns regarding this frequent and invasive intervention in this population.

This proof-of-concept protocol describes the evaluation of the safety, effectiveness and acceptability of novel PIVD fixation splint in pediatrics: the FIX I/V splint. Manufactured in Quebec, these splints are made of colored antimicrobial plastic that moulds to the arm or hand to enhance safety and reduce the reliance on gauze bandages and adhesive tapes. They are sweat-resistant, antibacterial, single-use, hypoallergenic and recyclable. Their application is straightforward, requiring no fabric bandaging, while facilitating continuous monitoring of the intravenous site. They are available in two models: one for the hand and another for the elbow, each coming in four sizes (small, medium, large, and extra-large), tailored to the child’s weight or age (Tremblay, 2023). Health Canada and the Food and Drug Administration (FDA) have approved the materials used in these splints (Czajkowski & al., 2015; Québec, 2026).

### Purpose, objectives, and hypotheses

This article presents the protocol for an interventional, prospective, non-randomized proof-of-concept study designed to evaluate the effectiveness, safety and acceptability of a novel pediatric splint, the FIX I/V splint. Specifically, the study pursues the following objectives:

1. Evaluate the effectiveness and safety of the FIX I/V pediatric fixation splint used on children aged between two weeks and eight years, 11 months, who require a PIVD near the wrist or elbow and are admitted to the pediatric units of two hospitals within an Integrated Health and Social services Center (CISSS) in Quebec, Canada (experimental group), and to compare these outcomes with those of the conventional fixation splint used on children of the same age group with a PIVD, admitted to the pediatric units of two hospitals within another CISSS in Quebec (control group).
  a. Effectiveness of the splint :
    i. The use of the FIX I/V splint shows a descriptive trend toward an increase in the lifespan of the PIVD compared to the control group wearing the conventional splint, thereby providing a preliminary signal of clinical effectiveness.
    ii. The use of the FIX I/V splint shows a descriptive trend toward a reduction in the number of PIVD reinsertions compared to the control group, thereby providing a preliminary signal of clinical effectiveness.
    iii. The use of the FIX I/V splint shows a descriptive trend toward a decreased incidence of PIVD-associated adverse events (such as infiltration, extravasation, phlebitis, local infection, occlusion or accidental removal) compared to the control group, thereby providing a preliminary signal of clinical effectiveness.
  b. Safety of the splint:
    i. The use of the FIX I/V splint shows a descriptive trend toward a reduction in device-related adverse events (such as skin injuries or pressure injuries) compared to the control group wearing the conventional splint, thereby providing a preliminary signal of clinical safety. An acceptable preliminary clinical safety signal will be defined as an adverse-event rate associated with the FIX I/V splint that is less than or equal to that of the conventional splint, thereby validating the feasibility and relevance of a larger-scale clinical trial.
2. Evaluate and compare the costs associated with nursing time spent on PIVD and splint installation, as well as routine monitoring activities, per patient and per shift, between the experimental group (FIX I/V splint) and the control group (conventional splint).
3. The use of the IX I/V splint shows a descriptive trend toward lower nursing-time costs related to PIVD and splint installation, as well as monitoring routines, per patient, per unit, and per shift, compared to those associated with the conventional splint.
4. Evaluate the acceptability of the FIX I/V fixation splint among healthcare staff and parents.
  a. Most of the healthcare staff and parents will express satisfaction with the safety of the FIX I/V splints, specifically: the splint characteristics, their child’s comfort, and their overall experience with the device.

## Méthodology

### Research design

To achieve the study objectives, a prospective, interventional, proof-of-concept (PoC) study is currently underway. This pilot approach (Eldridge & al., 2016; Thabane et al., 2010) serve a dual purposes. First, it fulfills an essential methodological objective: identifying a preliminary signal of extended lifespan for PIVD secured with FIX I/V splint. Second, it addresses an ethical imperative by initially limiting exposure to a small sample (N=50) to confirm the splint’s safety. The descriptive data generated through this approach will provide a rigorous scientific foundation for planning and optimizing a future, larger-scale randomized controlled trial. To ensure its methodological validity, the design of this study was guided by the SPIRIT recommendations for clinical trial protocol writing (Chan et al., 2013).

### Study context

This study was conducted in four pediatric units at two healthcare institutions in Quebec, involving hospitalized children who required PIVD placement between fall 2023 and winter 2024. Among these units, two had already implemented FIX I/V splints for PIVD fixation across their pediatric, emergency, and surgical departments, while the remaining two units used conventional splints. The research team presented the project to the assistant directors of nursing, senior advisors for continuous improvement, and the heads of the targeted units. These meetings, held remotely via Zoom, confirmed both the project’s feasibility and the care units’ interest in participating. Finally, pediatric nurses were informed about the study through an official letter distributed by their respective unit heads.

### Eligibility criteria

To be eligible for participation in the study, children had to be aged between two weeks and eight years and eleven months (regardless of ethnic origin or language). Inclusion required the administration of a prescribed intravenous treatment necessitating both PIVD placement and splint fixation. Splinting was indicated when the PIVD insertion site was located near the wrist or the elbow crease, or when the child exhibited agitation or unpredictable mobility. Patients meeting any of the following criteria were excluded from the study: 1) clinical instability (clinically unstable children), 2) major congenital anomalies, 3) terminal health conditions (end-of-life care), 4) compromised skin integrity precluding splint placement and 5) parental crisis situations (parents experiencing acute crisis).

### Outcome criteria

Adverse events related to PIVD include occlusions, infiltrations, extravasations, phlebitis, infections, nerve injuries, damaged catheter, air embolism, and deep vein thrombosis. The management and documentation of these complications must align with the Infusion Nurses Society (INS) infusion therapy standards of practice (Nickel et al., 2024; Nickel et al., 2026). Furthermore, PIVD fixation mechanisms must not restrict blood circulation or compromise skin integrity (Nickel et al., 2024; Nickel et al., 2026). Stabilizing devices must also be acceptable to both the child and their parents, allow unobstructed visual assessment of the catheter insertion site, and ensure no interference with the prescribed infusion flow rate (Nickel et al., 2024).

### Interventions

Experimental group. According to the FIX I/V splint user guide (Tremblay, 2023), the device can secure a PIVD placed either on the hand or at the elbow crease. There are four sizes for each model: small for ages 0 - 4 months (3 - 6 kg), medium for ages 4 - 18 months (6 - 10 kg), large for ages 18 months to 4 years (10 - 16 kg), and extra-large for ages 4 - 8 years (16 - 27 kg). The hand splint stabilizes the wrist while preserving finger mobility (see Figure 2). Once the target vein is identified, the PIVD is inserted and secured with a transparent dressing, after which the hand splint is applied. It is then secured with a Hypafix tape strip and reinforced with transparent adhesive film at both the wrist and the level of the proximal phalanges. The elbow splint is gutter-shaped to optimally fit the curvature of the arm, requiring two Hypafix tape strips and two transparent adhesive film bands (see Figure 1). For additional training resources, instructional video tutorials are accessible at the following link: https://fix-iv.com/education/.

**Figure 1.**
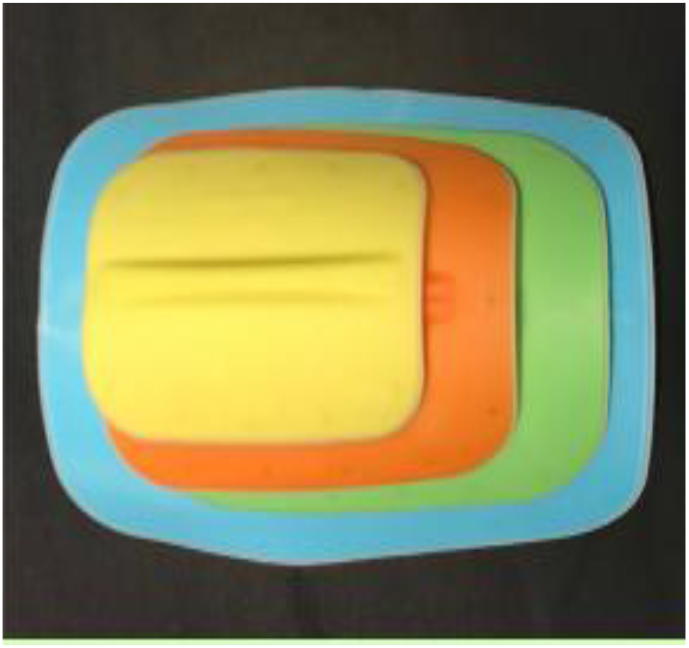

**Figure 2.**
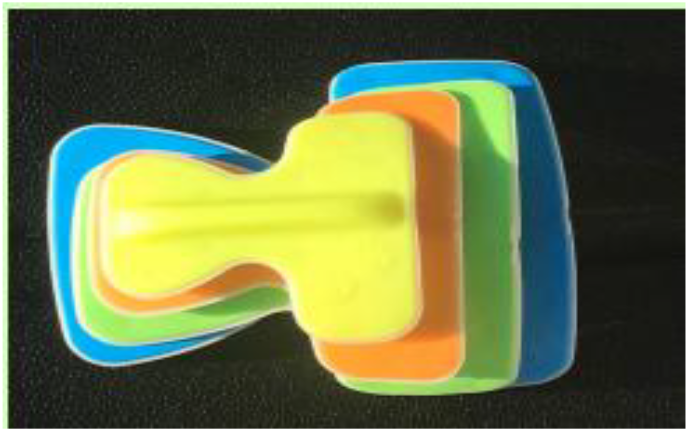
https://fix-iv.com/

Control group. The control group care team used conventional splints consisting of metal or plastic boards wrapped in fabric bandages. Once positioned, Hypafix bands and transparent adhesives secured the PIVD and immobilized the hand or arm to limit the risk of accidental removal. Participants in this group received standard care in accordance with institutional guidelines.

For both study groups, healthcare staff were required to assess and record the PIVD dwell time (in hours), the number of device reinsertions, catheter-associated adverse events (such as solution infiltration or extravasation, local infection, phlebitis, occlusion, or accidental removal) and the risk or occurrence of splint-induced pressure injuries. All four participating units followed uniform clinical guidelines for PIVD assessment, monitoring, and placement. In accordance with pediatric care standards, the intravenous site was monitored hourly. The protocol preparation timeline is detailed in Appendix Table 1.

**Table 1.** Protocol preparation timeline.

| October 2022 | Request from the FIX I/V company to evaluate their product in pediatrics and neonatology |
|---|---|
| November 2022 | Development of the MITACS-Accelerate grant application |
| August 2023 | Approval of the application and award of the \$30,000 grant |
| August 2023 | Drafting of the proof-of-concept protocol for ethical certification |
| September2023 | Hiring of two interns for the winter 2024 term |
| October 2023 | Conditional approval with minor modifications from the Central Research Ethics Committee of the Ministry of Health and Social services |
| November 2023 | Protocol approval by the ethics committee |
| November 2023 | Submission of the protocol for institutional suitability review to the experimental group's ethics committee |
| November 2023 | Study approval by the experimental group's ethics committee |
| January2024 | Launch of data collection for the experimental group |
| April 2024 | Institutional suitability approval granted for the control group |
| January 2025 | Launch of data collection for the control group |

### Sample size

Given that this is an exploratory, comparative, proof-of-concept study, the primary objective is not to achieve confirmatory statistical power, but rather to detect a preliminary descriptive signal of efficacy and to assess the acceptability of the FIX I/V splints. Consequently, the sample size was not determined by a traditional power calculation. Instead, a convenience sample consisting of 31 patients in the experimental group (FIX I/V) and 21 patients in the control group (conventional) was selected. This approach aligns with methodological recommendations by Julious (2005) and Whitehead et al. (2016) for pilot studies, which suggest a minimum of 12 to 15 patients per group to reliably estimate the variance of a continuous outcome, such as device lifespan in hours. The accrued sample (N = 52) is therefore sufficient to identify a descriptive trend and generate the necessary data to calculate effect sizes, with the goal of planning a future, larger-scale randomized clinical trial.

### Recruitment

Participants were recruited using a consecutive sampling method. Parents of eligible children were informed that a study evaluating fixation splints was being conducted. The research assistant (for the experimental group) and the assistant head nurse (for the control group) provided all necessary information regarding the study. They subsequently obtained free and informed consent from the parents to collect data both at the child’s bedside and from their electronic medical records. The detailed recruitment and data collection timeline is presented in Appendix Table 2.

**Table 2.**
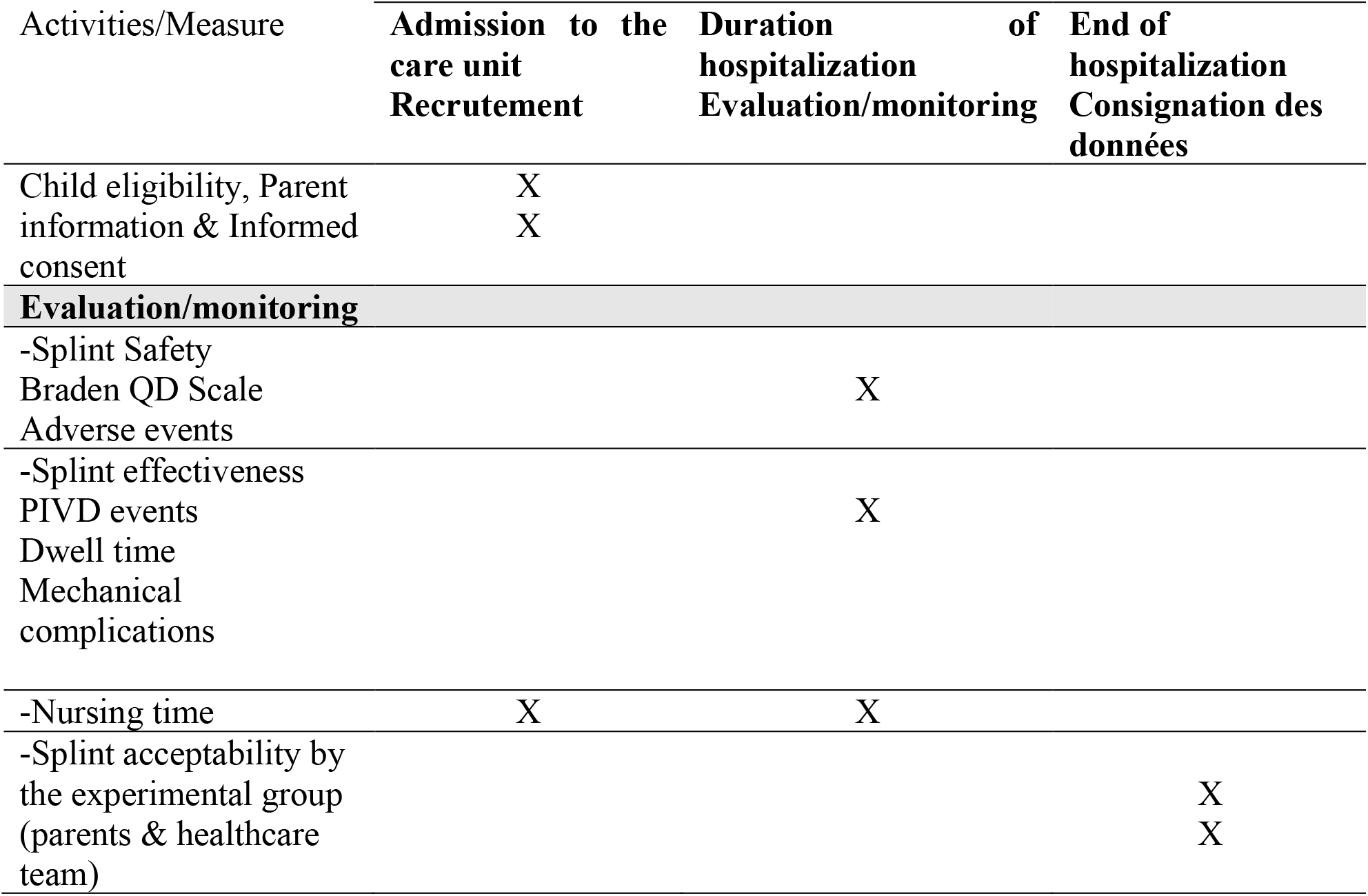
Recruitment and data collection timeline.

## Variables and data collection instruments

### Sociodemographic variables

The study collected sociodemographic data regarding the child (age in months at admission, ethnic group), the parent (age, country of birth, ethnic group, education level, professional status) and the participating nurse (age, education level, years of experience in pediatrics).

### Clinical variables

Clinical variables regarding the child (including admission date, weight at admission, primary reason for hospitalization, types of fluids and/or medications administered through the PIVD and date of discharge or transfer) were collected and recorded using a standardized data collection form integrated into the patient’s medical record.

### Splint effectiveness

Data on PIVC gauge, dwell time (in hours) and the number of reinsertions were recorded in a bedside data collection sheet.

### Splint safety

Adverse events associated with the PIVD (infiltration/extravasation, phlebitis, local infection, occlusion, or accidental removal) and the splint (moisture, erythema, skin maceration, or blistering) were recorded on the same bedside data sheet.

i. **Risk of splint related pressure injuries** Splint-related pressure injury risk was assessed using the pediatric Braden QD Scale (Curley et al., 2018). This tool predicts both immobility and medical device-related pressure injury risks in pediatric patients. It was validated in a cohort of 625 hospitalized children - ranging from preterm infants to individuals up to 21 years old - with a minimum stay of 24 hours and at least one medical device secured to the skin or mucous membranes. The scale evaluates seven criteria: 1) mobility, 2) sensory perception, 3) friction and shear, 4) nutrition, 5) tissue perfusion and oxygenation, 6) number of medical devices, and 7) device repositioning and skin protection. The first five criteria are scored from 0 to 3. For the remaining criteria, 1 point is assigned per medical device (up to 8 points), while device repositioning and skin protection are scored as 1 (potential problem) or 2 (problematic). A total cutoff score of 13 or higher demonstrates a sensitivity of 0.86 and a specificity of 0.59 in predicting pressure injury development (Curley et al., 2018).
ii. **Acceptability of FIX I/V splints** The acceptability of the FIX I/V splint among nursing staff and parents was evaluated using dedicated questionnaires. Staff acceptability was determined by assessing the splint’s ease of use and perceived effectiveness in maintaining PIVD patency for the intended duration (Feely et al., 2009; Harris et al., 2022). Conversely, parental acceptability was measured through overall satisfaction and their perception of the splint’s utility in reducing the need for adhesive tapes (Feely et al., 2009; Harris et al., 2022). Specific acceptability indicators are detailed in Tables 3 and 4.

**Table 3.**
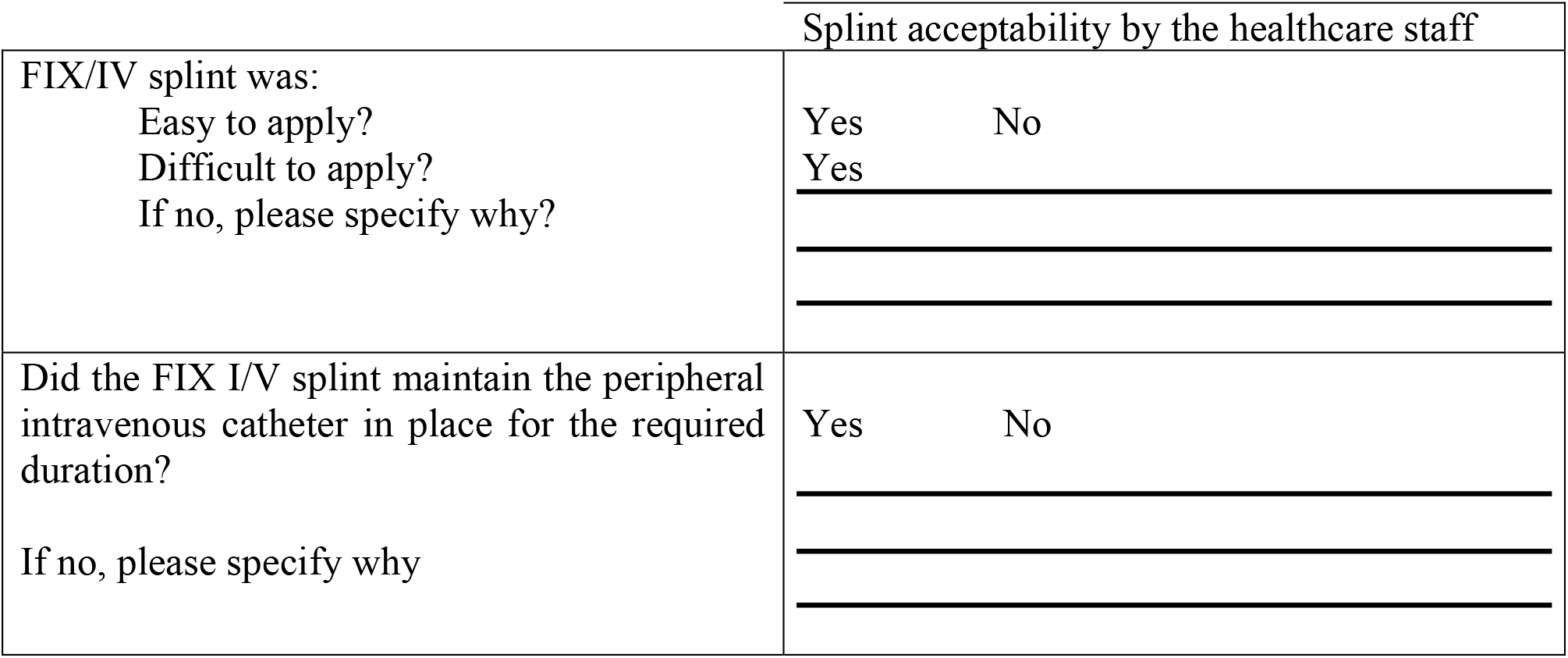
Staff specific FIX I/V acceptability indicators.

|  | Splint acceptability by the healthcare staff |
|---|---|
| FIX/IV splint was:<br>Easy to apply?<br>Difficult to apply?<br>If no, please specify why? | Yes No<br>Yes |
| Did the FIX I/V splint maintain the peripheral intravenous catheter in place for the required duration?<br>If no, please specify why | Yes No |

**Table 4.**
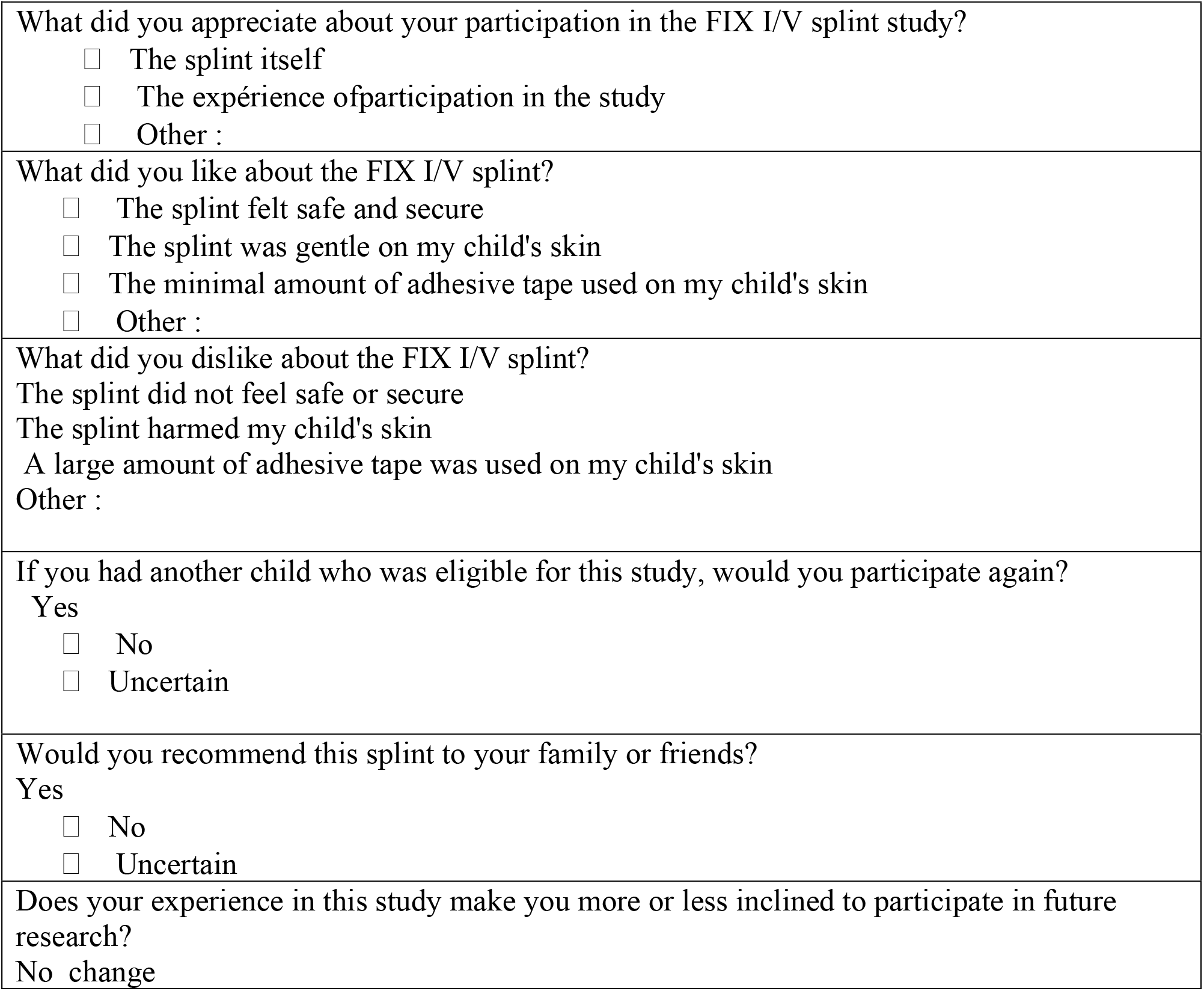
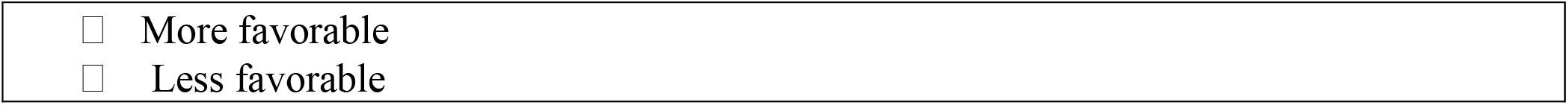
Parents specific FIX I/V acceptability indicators.

| What did you appreciate about your participation in the FIX I/V splint study?<br>The splint itself<br>The expérience ofparticipation in the study<br>Other : |
|---|
| What did you like about the FIX I/V splint?<br>The splint felt safe and secure<br>The splint was gentle on my child's skin<br>The minimal amount of adhesive tape used on my child's skin<br>Other : |
| What did you dislike about the FIX I/V splint?<br>The splint did not feel safe or secure<br>The splint harmed my child's skin<br>A large amount of adhesive tape was used on my child's skin<br>Other : |
| If you had another child who was eligible for this study, would you participate again?<br>Yes<br>No<br>Uncertain |
| Would you recommend this splint to your family or friends?<br>Yes<br>No<br>Uncertain |
| Does your experience in this study make you more or less inclined to participate in future research?<br>No change<br>More favorable<br>Less favorable |

### Data management and statistical methods

IBM SPSS Statistics (version 32.0) will be used for data entry and analysis. Due to the pilot nature of this comparative proof-of-concept study, analyses will be strictly descriptive and exploratory; no hypothesis testing or statistical significance analysis (p-values) will be performed. Continuous variables, notably PIVD lifespan (in hours), will be expressed as means with standard deviations or as medians with interquartile ranges, depending on distributional symmetry. Precision of measurement will be assessed using 95% confidence intervals (95% CI) to identify descriptive trends between the experimental group (n = 32) and the control group (n = 21). Categorical variables, such as occurrence of adverse events, will be presented as frequencies and percentages. Finally, the effect size (Hedges’ g) will be calculated from the lifespan data to quantify the magnitude of the clinical signal and to inform power and sample size calculations for a future confirmatory trial.

### Data monitoring and management of adverse events

Adverse event monitoring and management were conducted by the healthcare team. In compliance with quality of care directives, a formal incident report must be filed in the patient’s record for any adverse event or accident related to a medical device. These events were documented both in the bedside data collection sheet and within the progress notes of the patient’s medical record.

### Ethical considerations

This project received ethical approval from the Central Research Ethics Committee of the Ministry of Health and Social services on November 8, 2023 (project number: CCER 22-23-03). In addition, the two participating integrated health and social services centres conducted institutional suitability analyses and granted authorizations to carry out the study on their premises. During the study, a protocol amendment was submitted and approved to address low participation rates among the care teams. This amendment introduced a financial incentive (a lump-sum payment for the teams) in recognition of their collaboration in data collection.

Participants for the experimental group were recruited by two research interns who screened children upon admission to the units. The interns approached the parents of eligible children to present the study and provide the information and informed consent form. Parental participation consisted of completing a questionnaire regarding the acceptability of both the FIX I/V fixation splints and the study protocol. For the control group, assistant nurses informed eligible families and distributed the consent materials. Parents were explicitly notified that they would receive no direct personal benefit from participating, but that their collaboration would contribute to improving the quality of pediatric care. The sole inconvenience identified for parents was the time required to complete the questionnaire. Finally, all participants were informed that their involvement was entirely voluntary and that they maintained the right to withdraw from the study at any time without consequence.

Data collection from children and their parents was entirely anonymized by the research interns and healthcare teams. No identifying information or names were recorded on the forms, ensuring that investigators remained blinded to the participants’ identities and contact details. All data sheets were assigned a unique participant code to protect data confidentiality. Hard-copy forms are secured in a locked filing cabinet within the office of one of the principal investigators (AEH). Coded data were entered and stored on a password-protected computer under the same investigator’s supervision, with secure backups hosted on the UQAT server. The steering committee will have exclusive access to the full de-identified dataset and will oversee the analysis, interpretation, and reporting of results. Finally, findings from this study will be presented at relevant scientific conferences and published in peer-reviewed journals.

## Data Availability

All data produced in the present study are available upon reasonable request to the authors.

https://fix-iv.com/

